# Domain expert evaluation of advanced visual computing solutions for the planning of left atrial appendage occluder interventions

**DOI:** 10.1101/2022.04.11.22273553

**Authors:** Jordi Mill, Helena Montoliu, Abdel H. Moustafa, Andy L. Olivares, Carlos Albors, Ainhoa Aguado, Elodie Medina, Mario Ceresa, Xavier Freixa, Dabit Arzamendi, Hubert Cochet, Oscar Camara

## Abstract

Advanced visual computing solutions and 3D printing are starting to move from the engineering and development stage to being integrated into clinical pipelines for training, planning and guidance of complex interventions. Commonly, clinicians make decisions based on the exploration of patient-specific medical images in 2D flat monitors using specialised software with standard multi-planar reconstruction (MPR) visualisation. The new generation of visual computing technologies such as 3D imaging, 3D printing, 3D advanced rendering, Virtual Reality and in-silico simulations from Virtual Physiological Human models, provide complementary ways to better understand the structure and function of the organs under study and improve and personalise clinical decisions. Cardiology is a medical field where new visual computing solutions are already having an impact in decisions such as the selection of the optimal therapy for a given patient. A good example is the role of emerging visualisation technologies to choose the most appropriate settings of a left atrial appendage occluder (LAAO) device that needs to be implanted in some patients with atrial fibrillation having contraindications to drug therapies. Clinicians need to select the type and size of the LAAO device to implant, as well as the location to be deployed. Usually, interventional cardiologists make these decisions after the analysis of patient-specific medical images in 2D flat monitors with MPR visualisation, before and during the procedure, obtain manual measurements characterising the cardiac anatomy of the patient to avoid adverse events after the implantation. In this paper we evaluate several advanced visual computing solutions such as web-based 3D imaging visualisation (VIDAA platform), Virtual Reality (VRIDAA platform) and computational fluid simulations and 3D printing for the planning of LAAO device implantations. Six physicians including three interventional and three imaging cardiologists, with different level of experience in LAAO, tested the different technologies in preoperative data of 5 patients to identify the usability, friendliness, limitations and requirements for clinical translation of each technology through a qualitative questionnaire. The obtained results demonstrate the potential impact of advanced visual computing solutions to improve the planning of LAAO interventions but also a need of unification of them in order to be able to be uses in a clinical environment.

## 1. Introduction

Computing solutions such as 3D imaging and insilico simulation visualisation, Virtual/Augmented Reality (VR/AR) and visual analytics, among others, have seen an outstanding progress over the last years. Advances and improved access to big and high-resolution data, hardware infrastructure (e.g. High Performance Computing Clusters, Graphical Processing Units), affordable devices (e.g., VR/AR glasses) and Open-Source codes thanks to the commitment to Open and Reproducible Science by researchers, have been made it possible. Biomedical applications are not an exception, with an unprecedented availability to medical datasets at different multi-scale and resolution levels. Therefore, some visual computing technologies are already disrupting traditional concepts of medical image exploration. Complex medical interventions are usually planned by exploring preoperative medical images, where advanced visual computing solutions in combination with technologies such as 3D printing could contribute to reduce potential complications.

Until recently, the visualisation and analysis of medical imaging data was always performed with visualisation tools, usually as 2D images and standard multi-planar reconstruction (MPR) visualisation viewed in 2D flat monitors. The most common situation is that these are the only tools that can be used during intervention since they are fully integrated with the acquisition systems in the operating room, with clinicians mentally integrating the 3D structure and functional information provided by multiple sources. Off-line image analysis can be performed with numerous tools including Open-Source software such as 3D Slicer Kikinis et al. (2014) or commercial solutions, usually tailored to specific imaging modalities and type of pathologies. Most of these imaging tools are standalone software but web-based frameworks with Cloud-based engines are becoming modern and more flexible alternatives.

In cardiology, patient-specific detailed information about the structure and function of the heart is key for medical training and for optimising and personalising clinical decisions involving diagnosis, treatment planning and post-therapeutic monitoring of patients. Specifically in structural heart disease, where trans-catheter interventions are becoming a less invasive alternative to open surgery. However, the field of view in trans-catheter interventions is limited, with the absence of a gold-standard open cavity surgical field depriving physicians of the opportunity for tactile feedback and visual confirmation of cardiac anatomy Wang et al. (2021). At this juncture, there is a significant gap in understanding the 3D (plus time) anatomical and physiological relationships in the heart Wang et al. (2018a) that visual computing solutions can help to bridge. Recent studies have reviewed the added value of advanced visualisation of cardiac data, including in specific applications such as in congenital heart disease Salavitabar et al. (2020); Goo et al. (2020), in structural heart disease Wang et al. (2021) or transcatheter mitral valve replacement Kohli et al. (2020).

The left atrial appendage occlusion (LAAO), with a device inserted into the heart in a trans-catheter intervention, is a recent efficient alternative for patients in atrial fibrillation (AF) with contraindications to drug therapy. The interventional cardiologists mainly use multi-modal images in different 2D views such as echocardiography and X-ray to decide the optimal type and size of the device, as well as the position where to implant it in a given heart. Nevertheless, the complexity and inter-patient variability of the LAA 3D anatomy can be unnoticed with 2D imaging modalities. Three-dimensional echocardiography and more recently high-resolution Computerised Tomography (CT) images are increasingly becoming available, providing spatial information of the LAA to optimise the implantation strategy and device selection before the intervention Saw et al. (2016).

Commercial solutions are available with standard MPR visualisation of the 3D imaging modalities (e.g., CT images) such as the 3mensio Structural Heart software (Pie Medical Imaging, Bilthoven, the Netherlands) Hascoet et al. (2018), which also includes advanced 3D photorealistic rendering of the heart for an advanced exploration of the LAA or Materialise Mimics (NV, Leuven, Belgium). Karagodin et al. Karagodin et al. (2020) recently demonstrated an improved delineation of cardiac structures, including the LAA, with a new tissue transparency trans-illumination tool when visualising 3D echocardiographic images, comparing to standard 3D rendering in a system developed by Philips (Andover, MA). Most of these imaging tools to explore 3D LAA anatomies are based on stand-alone software but webbased frameworks with cloud-based engines are becoming modern and more flexible alternatives for medical image visualisation and analysis. The Virtual Implantation and Device selection in left Atrial Appendages (VIDAA) platform was recently developed, providing a clinician-friendly web-based tool to support the pre-operative planning of LAAO interventions Aguado et al. (2018). It allows clinicians to interactively explore the LA geometry as a 3D mesh with different Computer-Aided Design (CAD) LAAO models, together with some morphological indices and the original CT images in MPR format. However, these software tools are still limited to visualise and analyse imaging data with the standard MPR, volume rendering and surface mesh views in 2D flat monitors, thus with limited degrees of freedom interaction and preventing a correct perception of the 3D nature of the studied anatomy (e.g. depth, scaling).

Three-dimensional printing can already be considered a tool routinely used in certain cardiology fields, especially when dealing with abnormal heart anatomies Vukicevic et al. (2017) such as in congenital heart disease and paediatric applications Forte et al. (2019), to provide the clinician a better understanding of 3D cardiac anatomy. The clinical translation of 3D printing has been made possible due to the reduction of printer costs with simple rigid plastic materials, although more sophisticated printers and flexible materials mimicking tissue properties are available at a higher cost. Adaptation of 3D printing in clinical care and procedural planning has already demonstrated a reduction in early-operator learning curve or in centres without previous experience on trans-catheter interventions Wang et al. (2021); Fan et al. (2019); Wang et al. (2018b); Eng and Wang (2018). Several studies have evaluated the added value of 3D printed models for training and planning of LAAO interventions (e.g., Bieliauskas et al. (2017)). However, Conti et al. Conti et al. (2019) recently compared 3D-printing recommendations and implanted devices, with an agreement of only 35%. Moreover, computational costs and time required for models to be printed with realistic materials are not negligible in the clinical workflow.

Although in an earlier phase of development and application in the biomedical field, there already exist proof-of-concept studies of using VR technologies for cardiac devices such as the pre-operative planning of trans-catheter closure of cardiac deficiencies such as ventricular septal Mendez et al. (2018) or sinus venous defects Tandon et al. (2019); Southworth et al. (2020). Nam et al. Nam et al. (2020) used new functionalities of the 3D-Slicer Open-Source software (i.e., link with VR headsets) to develop a tool for the virtual testing, selection and placement of trans-catheter device closures of atrial and ventricular septal defects. Narang et al. Narang et al. (2020) recently demonstrated a reduction in measurement variability and time required when exploring 3D echocardiography and CT images in different cardiac pathologies, with users reporting easy manipulation of VR models, diagnostic quality visualisation of the anatomy and high confidence in the measurements. As for LAAO devices, the EchoPixel True 3D Virtual Reality Solution (EchoPixel, Inc., Mountain View, California, United States) allows to visualise CT scans and perform a “device-in-anatomy” simulation for LAAO preprocedural planning Sanon and Lim (2019). Zbronski et al. Zbroński et al. (2018) also visualised CT-derived LA anatomies before and during the occlusion procedure with the AR Hololens headset, being a useful enhancement according to clinicians. Finally, Medina et al. Medina et al. (2020) developed a VR-based platform (VRIDAA) for the visualisation/analysis of LAA anatomies and the most appropriate occlusion devices to be implanted, being positively evaluated by clinical users as a source of motivation for trainees and to better understand the required approach before the intervention. Nevertheless, the associated costs of high-end VR headsets and the spatial requirements can still be important barriers for the use of VR technology in a clinical environment. In-silico simulations from Virtual Physiological models of the heart, also known as Digital Twins, are emerging as a valuable technology in cardiology to support clinical decisions such as interventional planning, diagnosis and device optimisation Ribeiro et al. (2020); Corral-Acero et al. (2020). For instance, the commercial software FEops HEARTguideTM (FEops nv, Gent, Belgium) provides patient-specific structural simulations of LAAO deployments with different device configurations Garot et al. (2020), but without user interaction and lacking functional information. Computational Fluid Dynamics (CFD) solvers compute flow and pressure throughout a well-defined geometrical domain and can provide useful functional information about blood flow patterns with high spatial resolution, currently unattainable with in-vivo imaging techniques. Different therapeutic scenarios can be in-silico tested with CFD before the intervention, thus reducing operation costs with enhanced efficiency Naci et al. (2019) and accelerating research and development understanding of fluid mechanics within device testing Wang et al. (2021). Regarding LAA applications, several researchers have run CFD simulations to study blood flow in the left atria Otani et al. (2016); García-Isla et al. (2018); Bosi et al. (2018); Masci et al. (2020), including after the implantation of LAAO to find the optimal configuration of device Aguado et al. (2018); Mill et al. (2020). However, comprehensive preoperative simulations may take between hours and days depending on the complexity of the anatomy and potential interactions between cardiac tissue and the blood flow to be modelled Wang et al. (2021), thus limiting its application in a clinical environment.

In this manuscript, we present the evaluation of several advanced visual computing solutions for the planning of LAAO interventions (i.e., web-based platform with 3D imaging visualisation, virtual reality, in-silico fluid simulations), together with 3D printing with standard and affordable materials. 3D imaging data from CT scans of five patients candidates for LAAO implantation were visualised with the different visual computing technologies and 3D printing by six domain knowledge experts (three interventional and three imaging cardiologists). During the practical session, they were asked to decide the LAAO device settings after using each technology for each patient-specific case and to fill in an usability questionnaire and some open questions to assess the adding value, limitations and requirements for clinical translation of each one of the evaluated technologies.

## 2. Methodology

Figure 1 shows a general overview of the evaluation pipeline followed in our study. The original 3D CT scans of five patients, acquired before the LAAO intervention, were segmented to derive a binary mask of the left atria, including the left atrial appendage. Subsequently, a 3D model in the form of a surface mesh was generated and introduced, together with the grey-level 3D scan if required, to the specific processing workflow of each one of the evaluated computing technologies (e.g., web-based 3D imaging, 3D printing, VR and in-silico simulations). The main question asked to the study participants was to decide which device to implant for each patient. The LAAO devices selected for this study were the Amplatzer Amulet (St. Jude Medical-Abbott, St. Paul, Minnesota, United States) and the Watchman FLX (Boston Scientific, Marlborough, Massachusetts, United States), with their different sizes available commercially. Therefore, the participants of the study tested all the technologies with their different available features (see Section 2.3). After each technology, participants chose a given device configuration and were asked to give a final decision at the very end on device type, size and position to implant. Subsequently a System Usability Scale (SUS) questionnaire Brooke (1996) as well as some open questions (see Section 2.4.6) were filled in by each physician, focusing on the implantation of the tested technologies at their hospitals.

**Figure 1:**
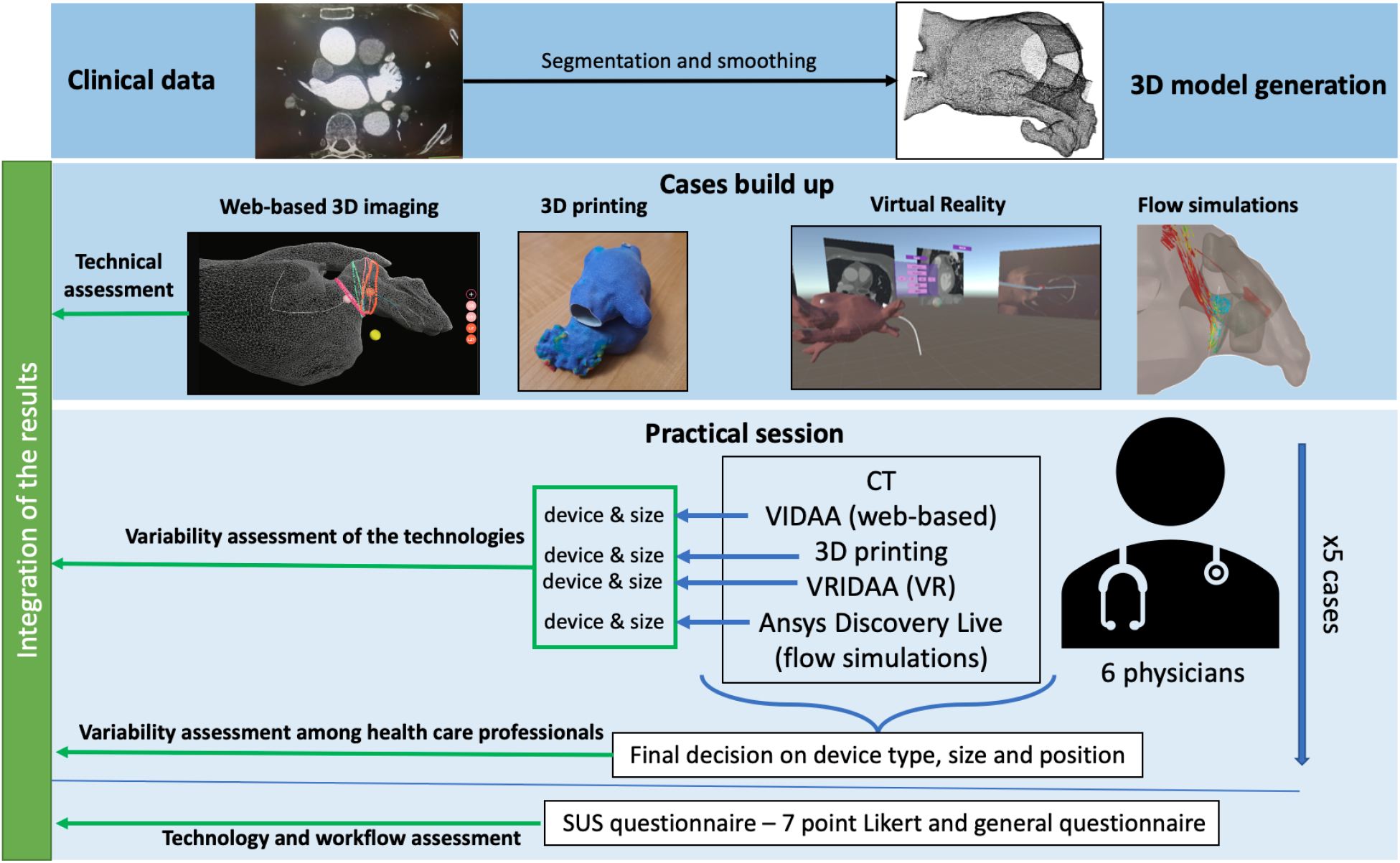
Overall pipeline for the evaluation of advanced computing technologies for the planning of left atrial appendage occlusion (LAAO) interventions. The first step involved generating the 3D surface model from the patient-specific medical images (i.e., Computerised tomography, CT) of five cases. The resulting 3D model was the base for the setup of all models used in the different technologies, which were tested by domain experts (i.e., physicians) in an experimental session where they needed to decide the device type, size and position. Subsequently, the participants answered a System Usability Scale (SUS) questionnaire and a general questionnaire with open questions.

### 2.1. Clinical data

The clinical data used in this work were provided by Hospital Haut-Lévéque (Bordeaux, France), including AF patients that underwent a LAAO intervention and with available pre-procedural high-quality CT scans. Five of them were randomly selected. The study was approved by the Institutional Ethics Committee; patients gave the informed consent for having their data used for research purposes, including tasks such as the ones presented in this study.

Cardiac CT studies were performed on a 64-slice dual source CT system (Siemens Definition, Siemens Medical Systems, Forchheim, Germany). Tube current was set to 120kV in patients with body mass index (BMI) > 27 and 100kV in those with BMI <27. Acquisition was set on end-systole using prospective ECG triggering, the delay being set in percentage of the RR interval in patients in sinus rhythm, and in ms in those with arrhythmia. Images were acquired using a biphasic injection protocol: 1mL/kg of Iomeprol 350mg/mL (Bracco, Milan, Italy) at the rate of 5mL/s, followed by a 1mL/kg flush of saline at the same rate. A bolus tracking method was applied to acquire arterial phase images, the region of interest being positioned within the LA.

### 2.2. 3D model generation

For each selected patient, the anatomy of the left atria, including its appendage, was extracted from the CT images using semi-automatic region-growing and thresholding tools available in 3D Slicer. The resulting binary mask of the LA was then introduced to the Marching Cubes algorithm to generate a 3D surface mesh model. Mesh smoothing was applied to correct irregularities from the segmentation, based on a Taubin filter smoothing operator (λ = 0.5, µ = − 0.53), followed by the removal of self-intersecting faces and non-manifold edges wherever necessary using Meshlab 2016.12 ^1^. The overall process of generating the 3D model took around 45 min per patient. In the following, a description of the set-up required for each evaluated computing technology is provided. Furthermore, Table 1 illustrates the estimated preparation times and associated costs for each technology.

**Table 1:** Set-up preparation (for each patient in brackets) and practical session times as well as the associated costs for each technology. The time required to build the 3D models, including medical image segmentation (3.75 hours) are not included. RP: research platform (i.e., no cost).

|  | Set-up preparation times (per case) | Average practical session duration per case | Cost |
| --- | --- | --- | --- |
| 3D printing | 43 h (26.1 h) | 3 min | 6,000 + 17.5 € |
| VIDAA | 7 min (1.24 min) | 8.13 min | RP |
| VRIDAA | 15 min (3 min) | 10 min | RP+1238€+2500 PC |
| In-silico simulations | 21.6 h (4.3 h) | 12 min | Free licence + 2,000 PC |

### 2.3. Computing technologies

#### 2.3.1. VIDAA: web-based exploration of 3D imaging

Both CT scans and the LA meshes were introduced to the VIDAA platform for visualisation (as MPR and 3D, respectively) and morphological analysis (see Figure 2). The 3D surface mesh of the LA can be visualised with different levels of transparency, both in solid and wire-frame formats. Some landmarks relevant to LAA interventions such as the circumflex artery, can be manually selected by the user. The centreline of the LAA was computed with the Python’s VMTK library^2^ from the centre of the LA mesh to a point on the LAA tip, the latter interactively selected by the user. Perpendicular 2D contours along the resulting centreline were then obtained to estimate morphological measurements on the LAA (i.e., maximum and minimum LAA diameters). These measurements were also visualised in 2D maps and a graph along the centreline depth to better identify local size variations in the LAA. The user can also define the ostium (i.e., intersection between the LA and LAA) and landing zone (i.e., where the device will be implanted) landmarks with small spheres along the centreline. Afterwards, based on the estimated measurements, the VIDAA platform proposes a set of appropriate LAAO devices for the studied LAA geometry. The user can interactively manipulate the chosen LAAO device, changing its position in the LAA and its size. Once the CT and the mesh were uploaded into the VIDAA platform, the case was ready to analyse, with the centreline and morphological measurement calculation taking less than a minute. Currently, the VIDAA platform is a research prototype and it is not available commercially.

**Figure 2:**
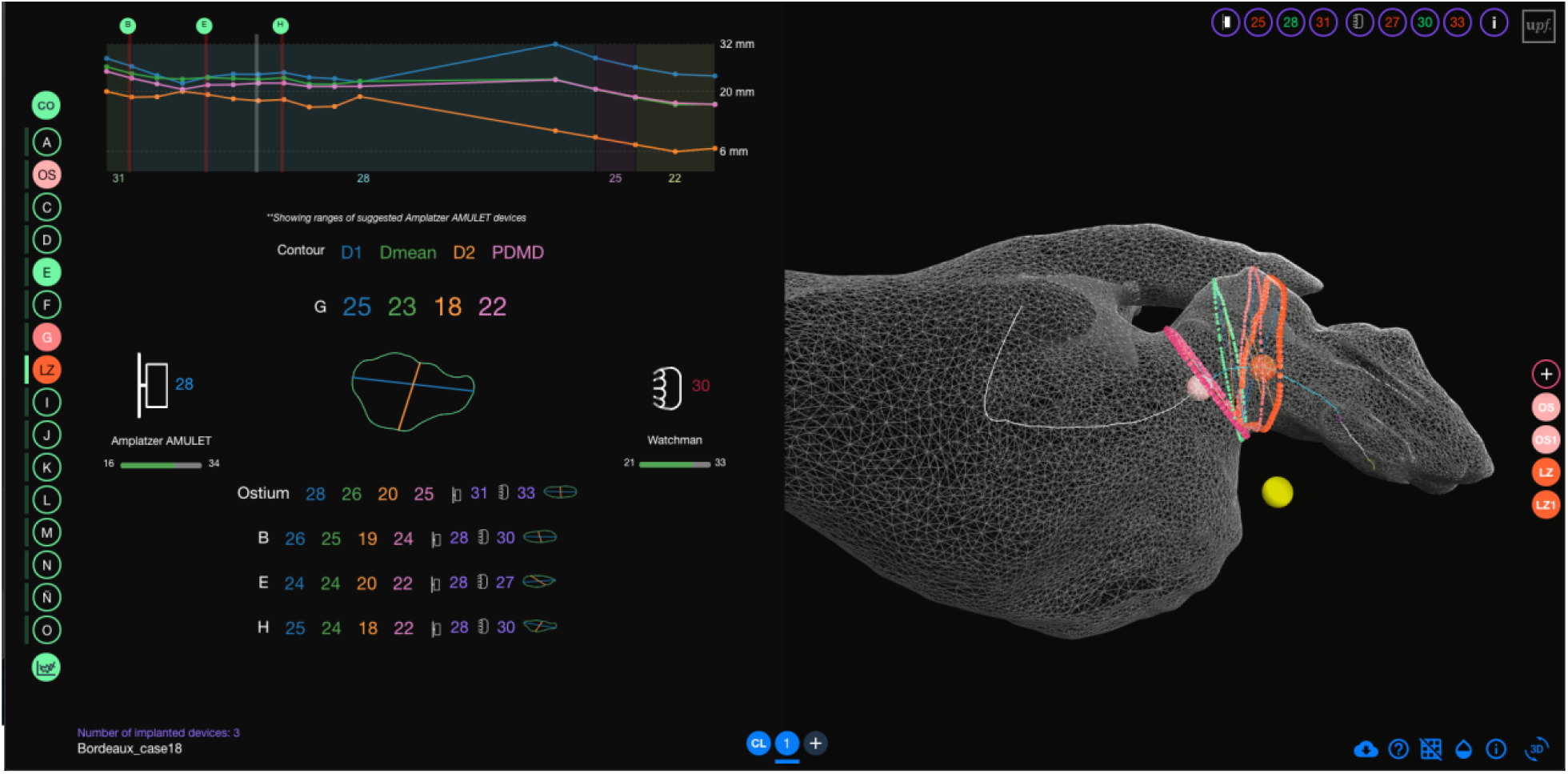
Web-based 3D imaging exploration (VIDAA platform). Left: Morphological parameters (e.g., diameters) of 2D contours along the centreline characterising the left atrial appendage (LAA) anatomy and range of most appropriate devices to implant. Right: 3D visualisation of the LAA anatomy in a 3D wire-frame mesh format, together with the LAA centreline (white), several 2D contours and some anatomical landmarks (pink, orange and yellow small spheres corresponding to the ostium, the landing zone and the circumflex artery, respectively) relevant for the device implantation.

#### 2.3.2. 3D printing

The 3D model generated from the CT scan of each patient was printed at Hospital de la Santa Creu i Sant Pau (Barcelona, Spain) with a Fused Deposition Modeling (FDM) 3D printer, the Ultimaker S5 (Ultimaker B.V.; Geldermalsen, Netherlands), which costs 6,000 euros approximately, depending on the vendor. The 3D LA model was prepared for printing with the Ultimaker Cura 3 software from the printer provider. Typical rigid Polylactic acid (PLA) was the material used for the LA models (see Figure 3), with an associated cost of 1.5 euros for each model. Twenty hours were needed to print all LA cases (i.e., 4 hours per case). Moreover, CAD models of Amplatzer and Watchman FLX LAAO devices corresponding to the different commercially available sizes were printed with thermoplastic polyurethane (TPU) to add more flexibility. Therefore, users could try to position the printed LAAO device into the 3D printed model of the LA to have more insight on their interaction. The cost of all printed LAAO devices was of 10 euros, taking 22 h to print. An extra hour was added for time estimations due to preand post-processing tasks such as adding thickness to the 3D models and removing the scaffolds of all the models.

**Figure 3:**
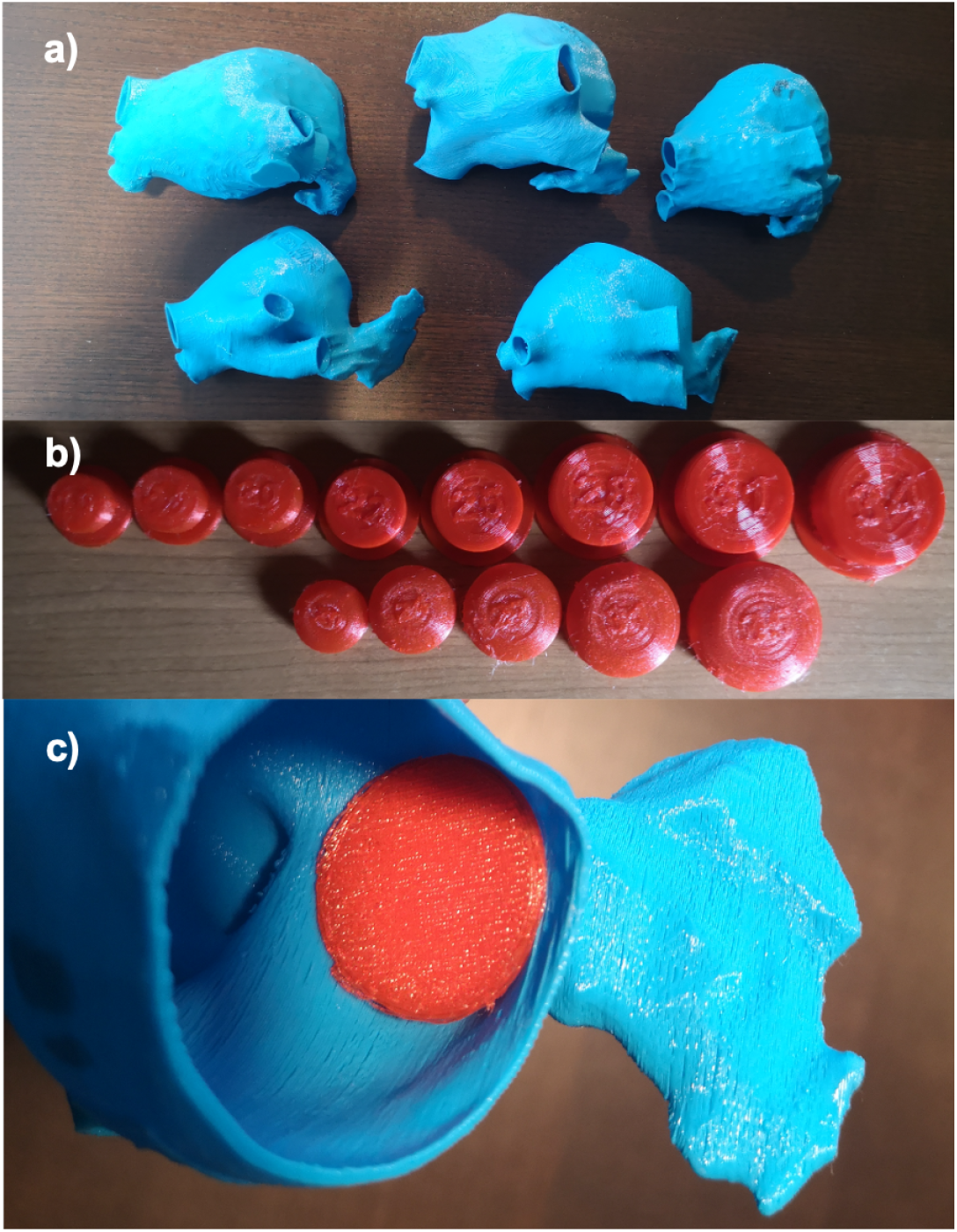
a) 3D printed models of left atria (LA) analysed in this study; b) 3D printed models of left atrial appendage occluder (LAAO) devices; c) Example of interaction between LA and LAAO 3D printed models.

#### 2.3.3. VRIDAA: virtual reality tool

The VRIDAA platform developed by Medina et al. Medina et al. (2020) was used to evaluate the use of VR technologies for the planning of LAAO interventions. It allows the user to interact with the LA geometry, jointly visualise it with patient-specific medical images in standard MPR format and relevant morphological indices. Standard surface manipulation including mesh clipping and transparency changes are possible, as well as browsing along the CT scan slices. Morphological measurements and landmarks imported from the web-based VIDAA platform such as the centreline and a graph with the associated LAA contour diameters can be also displayed in the VR environment. It is also possible to virtually place the LAAO device of choice (i.e., different designs, sizes) in any position. Additionally, the user can also plan the optimal location for introducing the delivery catheter into the LAA, freely manipulating a catheter model, together with an endoscopic view to facilitate the visualisation of the LA interior.

The computer used in this study was equipped with an Intel Core i5-8400 CPU @2.80 GHz processor, an NVIDIA GeForce RTX 2080 Ti graphic card and 16 GB of RAM, costing around 2,000 euros. For the implementation of the VRIDAA platform, the Unity Engine version 2018.1.8f1 (64-bit) (Unity Technologies, San Francisco, California, United States) was used, with SteamVR as the run-time and OpenVR as the API to get full compatibility with all major VR display platforms. The employed VR headset was the HTC Vive Pro, with a resolution of 1440 × 1600 pixels for each eye, a 90 Hz refresh rate, and 110 degrees field of view. Its cost was of 1,239 euros. Image processing outside the platform was performed using Python libraries. Once uploaded into VRIDAA, the user can interact with the 3D meshes by using the HTC Vive Pro controller to freely move it (6 degrees of freedom, i.e., rotations and translations) or zoom it in order to navigate inside the patient’s LA. In its current implementation, the VRIDAA platform is intended to be used with the possibility of moving within the virtual environment. Thus, a clear space of around 2 m x 1.5 m is required, as can be seen in Figure 4.

**Figure 4:**
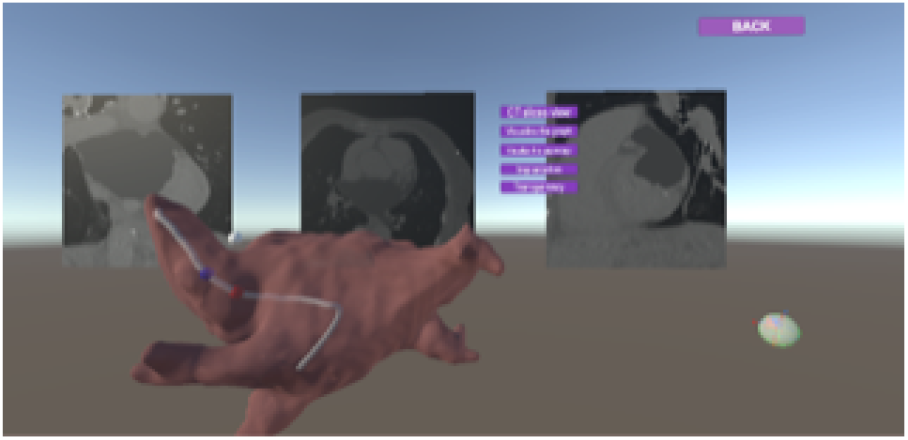
Virtual reality VRIDAA platform to explore left atrial (LA) anatomies and occluder devices. A 3D LA geometry with axial, sagittal and coronal slices of medical images visualised behind, together with the delivery catheter model (in white) and the 2D endoscopic view of the catheter’s tip camera for LAA interior visualisation.

All meshes and images were uploaded to VRIDAA before the practical session with participants. In VRIDAA, the centreline was not selected by the user; each case had a centreline previously generated in the web-based VIDAA platform, as well as the measurements and the proposed devices, which were transferred into the VR platform. Thus, if the system is already calibrated, the only required step for using the VR platform was the transfer and upload of the files to the VRIDAA system, making the workflow quite fast and straightforward.

#### 2.3.4. In-silico fluid simulations

The participants were presented with the visualisation of in-silico fluid simulations in the studied LA anatomies and including any possible LAAO device to be implanted, using the Ansys Discovery Live (ANSYS Inc, USA) software, under the Academic License (i.e., free of charge). It includes a GPU-based Lattice Boltzmann method that allows almost real-time simulations, once each case was set-up. Therefore, the participants could manually choose a given device (i.e., type, size) and place it in a given position to analyse resulting blood flow patterns, iterating as many times as desired.

The volumetric meshes required for the fluid simulations were generated from the 3D surface models using the Gmsh 4.5.4 software ^3^. The final meshes were around 800 thousand elements for all cases. The PC used to run the simulations was the one used for VR. The blood flow was treated as a Newtonian fluid, with a density of 1060 Kg/*m*^3^ and a viscosity of 0.0035 Pa/s. The boundary conditions in the inlets and outlets of the 3D LA model of the LA were chosen as in our most recent LAA fluid simulation study Mill et al. (2019), but without dynamic movement of the mitral ring plane since it was not allowed in the employed software. Basically, they were the same for all patients, extracted from Doppler echo and pressure measurements of an AF patient different from the ones processed in this study: a velocity profile in the outlet (i.e., the mitral valve) and a pressure wave at the pulmonary veins. The set-up for each simulation case included 30 min for meshing and building, accounting for 21.6 hours in total (with 13 devices per 5 patients).

### 2.4. Practical session

Before the practical session, the participants received a short tutorial on the different technologies, demonstrating their features. During the practical session data from 5 patients were presented to the physicians. For each case, the participants chose the optimal device settings after testing each technology. At the end of the case analysis, they were asked to make a final decision with all knowledge gathered from all technologies. Finally, they answered the SUS questionnaire and open questions for a global assessment of each employed technology.

The order to present the technologies in the practical session was the following: CT medical images (optional), VIDAA, 3D printing, VRIDAA and simulations. Initially, we offered the possibility of visualising the raw CT medical images of each patient. Afterwards, the web-based VIDAA platform followed, since it is similar to some of the most advanced software solutions commercially available in the market (e.g., 3mensio). Next, 3D printing and VRIDAA were presented to incorporate data visualisation with enhanced depth perception, starting with 3D printing since most physicians are more familiar with it. Finally, the in-silico simulations were shown in the Ansys Discovery Live software to include functional information complementary to the structural knowledge provided by the other technologies.

#### 2.4.1. CT imaging

The visualisation of CT medical images was optional since some of the physicians do not work on their daily basis with CT measurements. Although it was not assessed in SUS questionnaire, CT visualisation was offered since it is the gold-standard technique to explore LA anatomy, thus useful to plan LAAO interventions. Moreover, there were questions related to CT use in the general questionnaire. Physicians could explore the CT medical images with the Open-Source ITK-Snap software ^4^ to: (1) browse through the CT slices for inspecting the LAA shape and other anatomical landmarks such as the circumflex or the ostium; and (2) take measurements of these anatomical landmarks.

#### 2.4.2. VIDAA

Initially, after opening the 3D LA geometry in the VIDAA platform, participants were asked to select a point to create the optimal LAA centreline. Then, the contour diameters perpendicular to the LAA centreline were automatically computed (less than 30 sec), as well as the selection of recommended LAAO devices. Next, participants interactively explored the LAA geometry, observing the contour diameters and looking for the best position and device for each case. In addition, physicians could select among the different (recommended or not) LAAO devices and move them freely to make a decision.

#### 2.4.3. 3D printing

The 3D printed model of the LA of each studied case was offered to the physician to manipulate with their hands, together with the full range of 3D printed replicas of the Amplatzer Amulet and Wathchman FLX LAAO devices available in the market. Physicians could then interact with both type of 3D printed models to decide which device would fit better each LA anatomy.

#### 2.4.4. VRIDAA

The tasks performed by the participants on the VRIDAA platform were very similar to VIDAA. Nevertheless, the LAA centreline was already provided by default. Once the participant wore the VR glasses, the LA appeared, together with its centreline, and the physician could move it, grab it or go inside to better explore the interior of the anatomical structure. Then, the user could select a given device type and size, which would be placed at the beginning of the centreline, being able to move it along. In addition, the LAAO device could also be grabbed and moved freely.

#### 2.4.5. In-silico fluid simulations

Initially, the participant was asked to select a device type and size, after which it could be freely placed in any location of the LAA using the interface of the Ansys Discovery Live software. The LA anatomy could also be moved it. Once a device position was chosen, the simulation was launched, requiring a couple of minutes to visualise the resulting blood flow patterns (see Figure 5).

**Figure 5:**
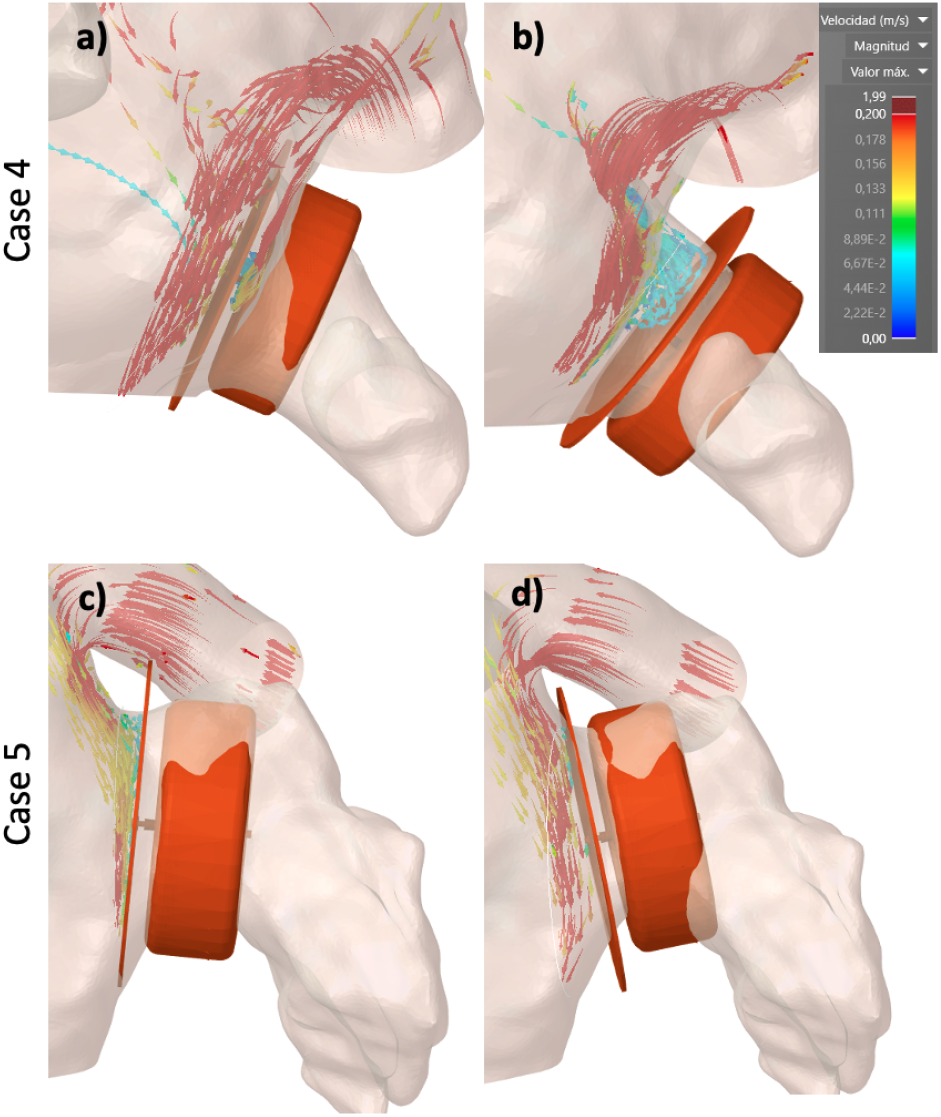
Examples of in-silico fluid simulations using Ansys Discovery Live with optimal device settings according to a given participant. 3D streamlines-based visualisation simulations were used to illustrate blood flow patterns of in the left atrium and left atrial appendage. Blue and red represent low velocity (<10 m/s) and high velocities (>20 m/s), respectively.

#### 2.4.6. Evaluation questionnaires

Once the participants had gone through all the patients, they answered two questionnaires: a SUS questionnaire and a more general questionnaire with open questions. The SUS questionnaire, developed by J.BrookBrooke (1996), was used to give a more quantitative assessment on the usability of the technologies. It is a ten-item questionnaire using, in our case, a 7-point Likert scale. Consequently, the physicians answered the SUS questionnaire for each technology at the end of the session. Details on the SUS questions and the answers for each technology are included as Supplementary Material.

The aim of the questionnaire with open questions was to profile the participants and to know more about how these technologies could be implemented in their current workflow, according to their point of view. The questions were the following:

- Years of experience in LAAO interventions.
- Current position at the hospital.
- Did you know about the application of these technologies to LAAO planning? If yes, which one?
- Have you tested any of these technologies before? If yes, which one?
- Have you participated in the development of these technologies? If yes, which one?
- Which technologies would you add in your ideal workflow for LAAO (disregarding economical and equipment restrictions)?
- Which technology did influence your final decision on device election the most?
- If your hospital is mainly using ultrasound imaging to plan LAAO interventions instead of CT, would you consider acquiring CT data only to be able to use these technologies?

## 3. Results

### 3.1. Participant profile

Out of the six participants three were interventional cardiologists (P2, P3, P4), i.e., physicians who are implanting the device, while the remaining three were imaging cardiologists, who are responsible for the medical image acquisition and analysis before and during LAAO procedures. In average, they had 5.08 years of experience in LAAO interventions (with 10 years and less than one for the most and less experienced, respectively). The participants work in three different hospitals; two of them using CT images for LAAO planning while ultrasound (US) imaging is the choice in the remaining clinical centre.

None of the participants have taken part in the development of VIDAA, VRIDAA or Ansys Discovery Live. However, one participant (P6) helped on the 3D printing process, but being blind to critical data in the study (i.e., which device was implanted, clinical output). Most participants (5/6) were familiar with 3D printing, having tested it before. Moreover, only one participant did not know about the use of any of these technologies for LAAO planning, while only one knew about all of them. Not a single participant had tested the VIDAA and VRIDAA platforms before the practical session and only two physicians had some experience with fluid simulations beforehand, although not with the Ansys Discovery Live software.

### 3.2. SUS questionnaire

The results of the SUS questionnaire are summarised in Table 2 and Figure 6. Overall, all the evaluated computing technologies passed the acceptability and user-friendliness threshold (as defined in Bangor et al. (2009)).

**Table 2:** SUS score for each technology and participant. I: imaging cardiologists; IC: interventional cardiologists. In bold, the best computing technology according to each participant.

| Participant | VIDAA | VRIDAA | 3D printing | Simulations |
| --- | --- | --- | --- | --- |
| P1 (I) | **88.0** | 70.4 | 72.0 | 51.2 |
| P2 (IC) | **72.0** | 62.4 | 64.0 | 70.4 |
| P3 (IC) | **67.2** | 59.2 | 64.0 | 28.8 |
| P4 (IC) | 72.0 | **80.0** | 78.4 | 73.6 |
| P5 (I) | **80.0** | 62.4 | 68.8 | 48.0 |
| P6 (I) | 89.6 | 75.2 | **92.8** | 77.6 |
| Mean (std) | 78.13 (9.24) | 68.2 (8.26) | 73.33 (10.96) | 58.27 (18.87) |

**Figure 6:**
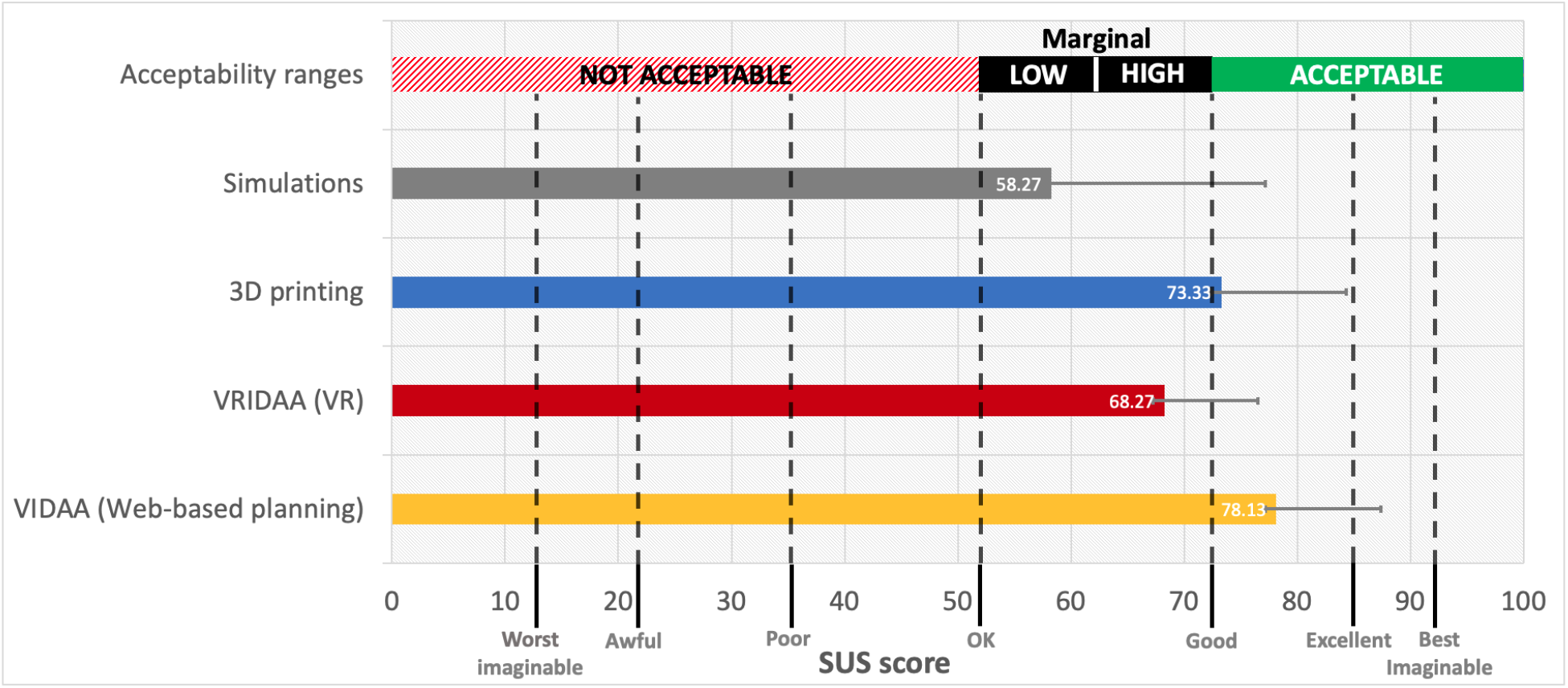
Overall results of the SUS questionnaire. Acceptable ranges were extracted from Bangor et al. (2009). Error bars show the standard deviation.

The web-based VIDAA platform was the best technology, according to the participants, with an average score of 78.13 and two physicians considering it excellent (scores above 85). Interestingly, as can be seen in Table 2, imaging cardiologists valued more VIDAA (average of 85.87) than interventional cardiologists (average of 70.4), although the latter still labelled the technology within the marginal high range of acceptability and a correct user-friendliness. The strongest points of the VIDAA platform, based on the SUS questions, were that it was easy-to-use and fast to learn without any support, with all participants agreeing on their willingness to use VIDAA frequently. On the other hand, the participants found that there were too many features and steps in the platform, which could be simplified, to perform the final LAAO planning.

3D printing was the second most valued technology (score of 73.3, i.e., with the acceptable range of usability), with good marks on easiness of usability and complexity. However, it failed on the confidence of use, consistency of the system and a proper integration of all features. Despite having 3D printed devices with flexible materials, being economic ones, the participants did not fully trusted them.

The virtual reality VRIDAA platform was the technology with a wider range of answers from the participants regarding its use on a daily basis, with one strongly agreeing to use it and the remaining ones with not clear opinion. Moreover, there was not agreement in the participants on any major flaw of the technology. Five participants consider it easy to use, although three of them reported that they might need support at some point. Participants also mentioned that the devices recommended by the VRIDAA system were slightly bigger than expected. Finally, according to the participants, most of them were confident with the device positioning due to the possibility of freely moving it.

The visualisation of in-silico fluid simulations was the technology with the lowest score (58.27), barely passing the usability test, in the low marginal area of acceptability ranges. However, it was the technology presenting the largest variance between participants (from 77.6 to 28.8), some evaluating it at the level of the remaining technologies and others in the not acceptable usability area. Also, five participants would like to use the technology frequently, with the remaining one providing an inconclusive answer. Moreover, they found most of the simulation-based features useful, in particular to see possible leaks after LAAO implantation, being well integrated and without inconsistencies, feeling quite confident on its use at the end of the practical session. The main reason for the overall low score was the poor easiness of use of the Ansys Discovery Live interface, almost all participants requiring support, and being difficult to learn and cumbersome to use.

### 3.3. Open questions

Figure 7 summarises the answers from participants to the open questions on each evaluated technology, focusing on their incorporation into the clinical workflow. Three physicians (all imaging cardiologists, not interventionists), would add the VIDAA platform into the clinical workflow, one of them only for planning complex anatomies and the two remaining one also including regular cases. The VR platform, VRIDAA, was including in the workflow by only 2/6 clinicians, both for regular and complex planning, while half of the participants would use 3D printing. However, 5/6 participants found the 3D printed models very useful for exploring the anatomy, willing to use this technology in a frequent basis, provided the cost of the printer and flexible materials would be cheaper. Despite the low values in the SUS questionnaire, 4/6 clinicians (including the 3 imaging ones) would use in-silico fluid simulations for planning complex anatomies, mainly to avoid leaks and device-related thrombus (DRT) after LAAO device implantation. It is worthy to point out that in-silico fluid simulations is the only technology positively rated for follow-up purposes (2/6 participants), especially if the relationship between low blood flow velocities and DRT is confirmed in more extensive clinical studies.

**Figure 7:**
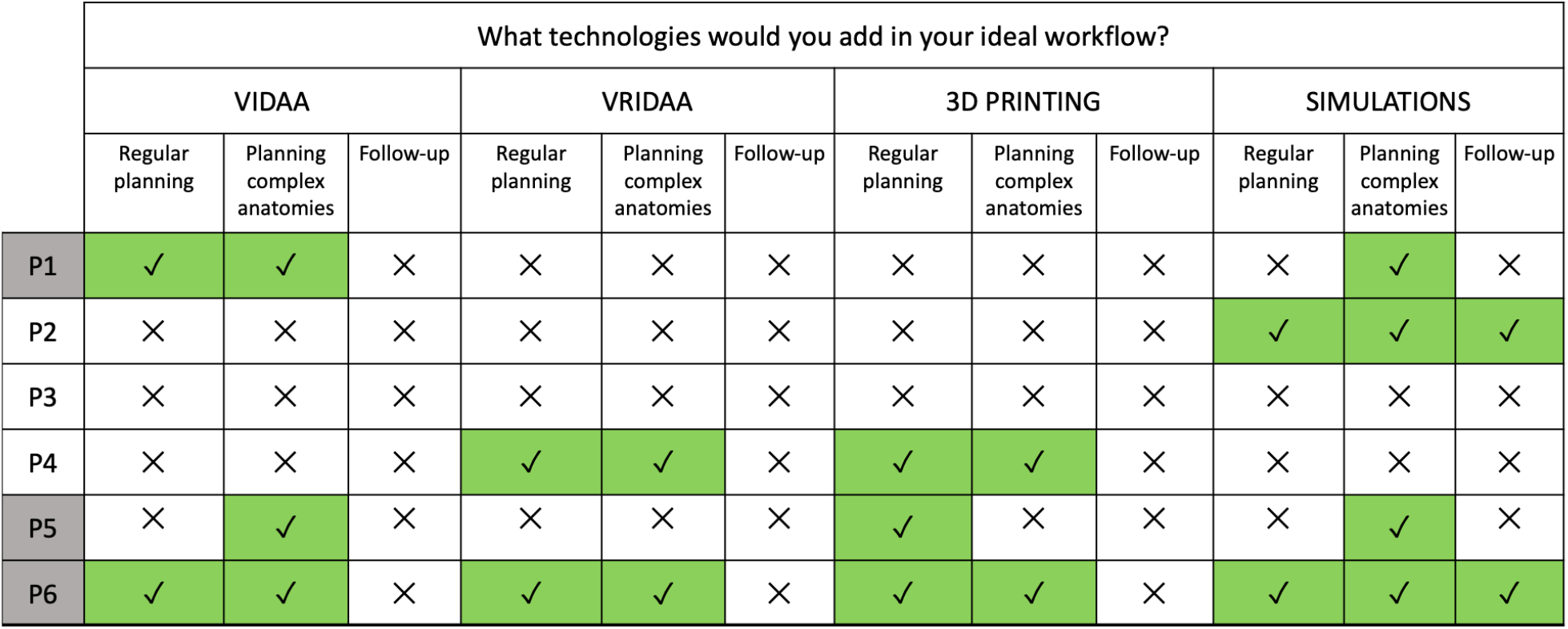
Ideal workflow according to the participants. Grey means imaging cardiologists, white interventional cardiologists.

One participant (P1) was from a hospital where CT is not routinely used for LAAO planning; ultrasound imaging is preferred since most patients are elderly people with other co-morbidities (e.g., renal dysfunction), often having contraindications to CT acquisition. However, this physician would be interested on acquiring CT scans to have access to the evaluated computing technologies when younger patients would be eligible for LAAO implantation.

Figure 7 also illustrates the extremes in physician approaches towards the use of these technologies, with one of them (P3) not willing to add any of them into the clinical workflow, and the counterpart one (P6) going for incorporating all of them. We would like to point out that P6 was the physician with more previous experience and interest on computational tools. An important remark from P6 was the need for a single software integrating the access to all different technologies.

### 3.4. Device selection comparison

Table 3 shows the final LAAO devices selected for each participant in all studied cases, compared to the device effectively implanted in the patient. Most LAAO devices selected by participants were the Amulet Amplatzer since it was the one they mainly saw in their training period and they had more experience with it. Therefore, they felt more comfortable using it.

**Table 3:** Devices finally selected by the participants. W: Watchman Flex; A: Amplatzer Amulet. Numbers refer to the device size (in mm). In bold, the cases that matched the size of the device implanted to the patient.

| Case n° | P1 (I) | P2 (IC) | P3 (IC) | P4 (IC) | P5 (I) | P6 (I) | Implanted |
| --- | --- | --- | --- | --- | --- | --- | --- |
| C1 | **A22** | A25 | **A22** | **A22** | A28 | **A22** | A22 |
| C2 | A18 | A18 | A22 | **A16** | A22 | A18 | A16 |
| C3 | A20 | A22 | A22 | **A18** | A22 | **A18** | A18 |
| C4 | A20 | W24 | A20 | A18 | A22 | A18 | A25 |
| C5 | A22 | **A25** | **A25** | **A25** | **A25** | W20 | A25 |

In three of the studied cases (C2, C3, C4), there were substantial intra-participant variations (e.g., more than 2 device sizes), while in the remaining cases, final decisions were quite similar. The main reason for these variations was the different LAA morphologies of the two groups of cases; the first group had a so-called chicken wing-type morphology that allows a different interventional technique (e.g., sandwich) with larger LAAO device sizes, which is preferred by some physicians. Unfortunately, the sandwich technique was not considered in any of the studied technologies. On the other hand, the agreement in the non-CW LAA cases (i.e., C1, C5) was higher, as can be seen in Table 3. Shockingly, none of the participants proposed the LAAO device finally implanted in case C4.

Table 4 illustrates the inter-participant variation in LAAO device selection after testing each computing technology, where different patterns can be observed. For instance, participants P1 and P5 (both imaging specialists) had a tendency to select smaller devices with 3D printing, which they attributed to the rigidity of LA walls in the printed model. Therefore, they mainly used the other technologies for their final LAAO device decision. Participants P2 and P4 (both interventional cardiologists) also followed the same pattern, without much LAAO size variation between different technologies. However, they were inclined to select a larger device in the VIDAA platform since it is not obvious to check for potential leaks in it, thus overestimating the size. On the other hand, features in the VRIDAA platform (e.g., being able to be within the LA cavity) and in-silico simulations (e.g., functional flow information), made these two technologies better suited for a more optimal device position to avoid leaks. Finally, participant P6 rarely changed the selected device after testing each technology.

**Table 4:** Devices selected by the participants (P1-P6) after using each computing technology. Web: web-based VIDAA platform; VR: virtual reality VRIDAA platform; 3Dpr: 3D printing; Sim: fluid simulations; W: Watchman Flex; A: Amplatzer Amulet. Numbers refer to the device size (in mm). In bold, when the device settings coincide with the final decision made by the clinician.

| Case | P1 (I)<br>Web/VR/3Dpr/Sim | P2 (IC)<br>Web/VR/3Dpr/Sim | P3 (IC)<br>Web/VR/3Dpr/Sim | P4 (IC)<br>Web/VR/3Dpr/Sim | P5 (I)<br>Web/VR/3Dpr/Sim | P6 (I)<br>Web/VR/3Dpr/Sim |
| --- | --- | --- | --- | --- | --- | --- |
| C1 | A25/A**22**/A16/A**22** | W24/W24/A25/A**22** | A28/A**22**/A20/A**22** | A25/A**22**/A**22**/A**22** | A25/A**28**/A20/A**28** | A**22**/A**22**/A**22**/A**22** |
| C2 | A20/A**18**/A16/A16 | A22/A**18**/W20/A**18** | A**22**/A**22**/A20/A**22** | A20/A**16**/A**16**/A**16** | A20/A**22**/A16/A**22** | A**18**/A**18**/A**18**/A**18** |
| C3 | A22/A20/A16/A**18** | W24/A**22**/W20/A**22** | A**22**/A**22**/A20/A**22** | A**18**/A**18**/A**18**/A**18** | A**18**/A**18**/A**18**/A**18** | A20/A**18**/A**18**/A**18** |
| C4 | A**20**/A**20**/A18/A18 | W27/W27/W20/W**24** | A22/A22/A**20**/A**20** | A20/A**18**/A**18**/A**18** | A20/A**22**/A18/A**22** | W20/A**18**/A**18**/A**18** |
| C5 | A25/A25/A18/A**22** | A28/A**25**/A22/A**25** | A28/A28/A**25**/A**25** | A**25**/A**25**/A**25**/A**25** | A**25**/A**25**/A20/A**25** | W24/W20/W20/W**20** |

## 4. Discussion

The fields of visual computing and 3D printing have seen a considerable progress over the last few years, slowly providing solutions for advanced visualisation in some biomedical applications. According to Dr. Wang, a well-known cardiology leader in the field of LAAO interventions, in a recent review Wang et al. (2021), there is a role for 3D printing, computational modelling and artificial intelligence (AI) to help bridge the dichotomy of realworld in-the-trenches imaging, and futuristic capabilities of computer science and biomedical engineering.

However, the clinical translation of advanced visualisation technologies is not straightforward, fulfilling the demanding requirements to be embedded in the existing workflows in hospitals. Despite recent generic reviews of visual computing solutions in cardiology applications (e.g., Wang et al. (2021); Salavitabar et al. (2020); Goo et al. (2020); Kohli et al. (2020)), there is a lack of complete studies testing the different visualisation methodologies on the same patient-specific data for benchmarking purposes. To the best of our knowledge, our study is the first attempt on this direction focused on LAAO interventions, aiming at evaluating the added value, limitations and requirements for the clinical translation of these technologies. The results obtained in the practical session demonstrated that the tested visual computing solutions are complementary, all providing added value in different steps of the current LAAO clinical workflow. All the evaluated technologies passed the threshold of acceptability range on usability, with the web-based VIDAA platform and 3D printing as the better rated, the former getting excellent marks by some participants. The VR-based platform (VRIDAA) and in-silico simulations were placed in the high and low marginal ranges, respectively, but the latter with large discrepancies between participants. Overall, the main conclusions among the participants was the complementarity of the technologies and the need for an integrative unique platform of the visual computing technologies (i.e., VIDAA, VRIDAA, fluid simulations) in order to be incorporated into the clinical workflow and used in a daily basis. Additionally, a more realistic elastic behaviour of the LAAO devices would increase the precision on the selected settings.

One of the most valued features in VIDAA was the detailed characterisation of the LAA anatomy, with the diameters along the centreline, since it can be used to identify the optimal implantation or better plan special strategies such as the sandwich technique. Additionally, based on the SUS questionnaire, VIDAA was fast and intuitive, with several manual steps but with a fast learning curve. Participants stressed the added value of the web-based platform in complex cases, proposing to incorporate functional information from fluid simulations for a more complete solution. The current commercial solutions comparable to VIDAA include stand-alone 3mensio Structural Heart software (Pie Medical Imaging, Bilthoven, the Netherlands),or Materialise Mimics (NV, Leuvren, Belgium) which include MPR visualisation, 3D rendering and 3D surface visualisation and FEops (FEOPSnv, Gent, Belgium), which provides simulations of device deployment. The VIDAA platform provides a more comprehensive and interactive morphological characterisation of the LAA, as well as interaction with 3D models of the LAA devices in a web-based environment that does not require any software installation and easily allows multi-centric studies and collaborative decisions. Moreover, none of these solutions offer in-silico fluid simulations. The price of these commercial softwares can also be an obstacle for including them in the clinical workflow of some hospitals.

Participants in our study acknowledged the better exploration of the 3D LAA anatomy with the VR system, due to an enhanced depth perception, 6 degrees of freedom interaction with 3D objects (both the LA geometry and the device) and views from the interior of the cavity (not easy to see even in 3D printed models), all points important for the device implantation Medina et al. (2020). For example, it was challenging for participants to truly grasp the depth and scaling of human organs and device sizes (as well as their relation) only from 2D monitors, specially to detect possible leaks. Although the learning times for using the VRIDAA platform were short (i.e., a few minutes), the participants preferred the combination of web-based 3D imaging software in conjunction with 3D printing since it would be easier to fit in the current clinical workflow. The evaluated VR setup, requiring a certain allocated space is not adequate for use in most hospitals. However, affordable VR headsets with reasonable performance and resolution, including wireless solutions without requiring much space (e.g., the Oculus brand or even augmented reality glasses) could represent a more appropriate alternative.

3D printing emerged as a useful technology for rapid prototyping, testing and pre-operative planning. However, the use of cheap materials in our study was a limiting factor since it did not realistically mimic the left atrial wall elastic properties, which is important to determine the interaction with the device once implanted. Specifically, it made physicians to usually pick smaller sizes than the one implanted or selected with the other technologies. The use of more realistic materials, such as the transparent and flexible HeartFlex from Materialise (NV, Lueven, Belgium) or resin, was also noted by the participants, but would dramatically increase the costs of the technology (approximately 200 euros per piece vs 1.5 euros with PLA). On the other hand, most physicians thought it was the best technology to recognise the shape and do a mental quick strategy of the intervention for regular planning.

In-silico fluid simulations including LAAO devices were a unique source of valuable functional information, not available from current imaging modalities or other computing solutions. Imaging cardiologists particularly valued this option for evaluating regions with potential leaks and complex flow after the LAAO implantation. However, the Ansys Discovery Live interface was not as user-friendly as the remaining technologies, as quantified in the SUS questionnaire, being difficult to interact with or move the CAD model of the device. Participants would not include this technology in its current form but they would recommend to incorporate in other tools such as VIDAA or VRIDAA.

The comparison between the devices selected by the participants after each technologies demonstrated the relevance for interventional cardiologists to explore the data fully in 3D with systems such as VR and including functional information from flow simulations. In addition, there were consistent differences in device sizes selected with 2D-based tools compared to 3D alternatives.

An additional interesting point risen by the physicians was the potential role of 3D printing and the virtual reality VRIDAA platform for training clinical fellows, especially on challenging cases. A complementary and very important use of these technologies according to domain experts involves patient education, which could contribute to reduce the levels of stress and anxiety prior to the intervention thanks to a better understanding of the procedure. A final comment was related to the application of the assessed visual computing solutions to other structural heart disease procedures involving medical devices such as trans-aortic valvular interventions since the technologies would only need slight modifications from the current LAAO-based use case.

There are some limitations in our study, starting by the small cohort of 6 participants that were able to evaluate the computing technologies. Moreover, all participants were trained with Amulet, thus undoubtedly creating some bias favouring such a device. Furthermore, the five studied cases were the same for all technologies. Therefore, participants remembered their choices with previously tested technologies, making their analysis not fully independent.

Additionally, we evaluated all computing technologies on 3D models built from CT medical images, which are not always available for LAAO planning. Echocardiography images could also be used in the presented visual computing solutions, mainly with advanced 3D rendering or in VR setups (e.g. Narang et al. (2020); Karagodin et al. (2020)), but anatomical details of the structures under study would be lost, which could negatively affect the medical decisions on the devices to implant. Otherwise, the manual steps and time required for creating the 3D models from the original medical images could be a limitation for the clinical translation of the evaluated tools. Nevertheless, the use of automatic deep-learning techniques for LA segmentation and mesh creation are being developed and should be available in the near future. Furthermore, it would have been useful to include new photorealism and advanced cinematic rendering visualisation Karagodin et al. (2020) in our study, which is planned in the future. Finally, an evaluation study where the advanced visual computing solutions are implemented and tested in the hospital would complete the current work, where technologies were tested by domain experts in a research lab.

## 5. Conclusions

In this work, we evaluated several computing technologies to assess their added value, limitations and requirements for being translated in a clinical environment, particularly for the planning of left atrial appendage occluder interventions. All the evaluated technologies could be beneficial in different steps of the LAAO clinical workflow, even if most need some adaptation to fit in the hospital. Specifically, the web-based 3D imaging VIDAA platform provided a complete morphological characterisation and excellent user-interaction to manipulate and test multiple device configurations. Economical 3D printed models, although lacking completely realistic device-LAA interaction, were useful to have a better perception of the 3D LAA anatomy and can easily be integrated in the current clinical workflows. VR technologies were also very helpful for 3D perception, being especially suited for educational or pre-operative planning purposes, but only simple VR headsets would be suitable for daily clinical routine. In-silico fluid simulations with LAAO devices resulted advantageous to reduce the risk of leaks and device-related thrombus after the implantation, but required more user-friendly interfaces. In consequence, all the evaluated computing technologies could contribute to better personalise LAA intervention and post-interventional treatment to each patient, ensuring better outcomes. It is not risky to foresee that advanced versions of the studied computing solutions will be properly embedded in clinical workflows in the near future, especially if they can be all integrated into a single space or software, as most participants of this study demanded.

## Supporting information

Supplemental SUS questionnaire

## Data Availability

All data produced in the present work are contained in the manuscript

## 6. Competing interests

The authors declare that they have no competing interests.

## 7. Acknowledgements

The authors of this manuscript want to thank the physicians, Dr L.S., Dr X.M., Dr L.A., Dr P.L. and Dr V.A., who accepted our invitation to take part in this study as participants. This work was supported by the Spanish Ministry of Science, Innovation and Universities under the Retos I+D (RTI2018-101193-B-I00), the Maria de Maeztu Units of Excellence (MDM-2015-0502), the Formation of Doctors (PRE2018-084062).Additionally, this work was supported by the H2020 EU SimCardioTest project (Digital transformation in Health and Care SC1DTH-06-2020; grant agreement No. 101016496).

## Footnotes

1 http://www.meshlab.net/

2 https://www.vmtk.com

3 https://gmsh.info/

4 http://www.itksnap.org/

## Notes

### Competing Interest Statement

The authors have declared no competing interest.

### Author Declarations

Ethics committee of Centre Hospitalier Universitaire Bordeaux gave ethical approval for this work

## References

Aguado, A.M., Olivares, A.L., Yague, C., Silva, E., Nunez-García, M., Fernandez, A., Mill, J., Genua, I., de Potter, T., Freixa, X., Camara, O., 2018. In silico optimization of left atrial appendage occluder implantation using interactive and modelling tools. accepted in Frontier: Cardiac Modeling: Aiming for Optimization of Therapy.

Bangor, A., Kortum, P., Miller, J., 2009. Determining what individual sus scores mean: Adding an adjective rating scale. Journal of usability studies 4, 114–123.

Bieliauskas, G., Otton, J., Chow, D.H., Sawaya, F.J., Kofoed, K.F., Søndergaard, L., De Backer, O., 2017. Use of 3-Dimensional Models to Optimize Pre-Procedural Planning of Percutaneous Left Atrial Appendage Closure. JACC: Cardiovascular Interventions 10, 1067–1070.

Bosi, G.M., Cook, A., Rai, R., Menezes, L.J., Schievano, S., Torii, R., Burriesci, G., 2018. Computational Fluid Dynamic Analysis of the Left Atrial Appendage to Predict Thrombosis Risk. Frontiers in Cardiovascular Medicine 5, 1–8.

Brooke, J., 1996. Sus: a “quick and dirty’usability. Usability evaluation in industry 189.

Conti, M., Marconi, S., Muscogiuri, G., Guglielmo, M., Baggiano, A., Italiano, G., Mancini, M.E., Auricchio, F., Andreini, D., Rabbat, M.G., Guaricci, A.I., Fassini, G., Gasperetti, A., Costa, F., Tondo, C., Maltagliati, A., Pepi, M., Pontone, G., 2019. Left atrial appendage closure guided by 3D computed tomography printing technology: A case control study. Journal of Cardiovascular Computed Tomography 13, 336–339.

Corral-Acero, J., Margara, F., Marciniak, M., Rodero, C., Loncaric, F., Feng, Y., Gilbert, A., Fernandes, J.F., Bukhari, H.A., Wajdan, A., Martinez, M.V., Santos, M.S., Shamohammdi, M., Luo, H., Westphal, P., Leeson, P., DiAchille, P., Gurev, V., Mayr, M., Geris, L., Pathmanathan, P., Morrison, T., Cornelussen, R., Prinzen, F., Delhaas, T., Doltra, A., Sitges, M., Vigmond, E.J., Zacur, E., Grau, V., Rodriguez, B., Remme, E.W., Niederer, S., Mortier, P., McLeod, K., Potse, M., Pueyo, E., Bueno-Orovio, A., Lamata, P., 2020. The ‘Digital Twin’ to enable the vision of precision cardiology. European Heart Journal.

Eng, M., Wang, D., 2018. Computed Tomography for Left Atrial Appendage Occlusion Case Planning. Interventional Cardiology Clinics 7.

Fan, Y., Yang, F., Cheung, G.S.H., Chan, A.K.Y., Wang, D.D., Lam, Y.Y., Chow, M.C.K., Leong, M.C.W., Kam, K.K.H., So, K.C.Y., Tse, G., Qiao, Z., He, B., Kwok, K.W., Lee, A.P.W., 2019. Device Sizing Guided by Echocardiography-Based Three-Dimensional Printing Is Associated with Superior Outcome after Percutaneous Left Atrial Appendage Occlusion. Journal of the American Society of Echocardiography : official publication of the American Society of Echocardiography 32, 708–719.e1.

Forte, M.N.V., Hussain, T., Roest, A., Gomez, G., Jongbloed, M., Simpson, J., Pushparajah, K., Byrne, N., Valverde, I., 2019. Living the heart in three dimensions: applications of 3D printing in CHD. Cardiology in the Young 29, 733–743.

García-Isla, G., Olivares, A.L., Silva, E., Nuñez-Garcia, M., Butakoff, C., Sanchez-Quintana, D., G. Morales, H., Freixa, X., Noailly, J., De Potter, T., Camara, O., 2018. Sensitivity analysis of geometrical parameters to study haemodynamics and thrombus formation in the left atrial appendage. International Journal for Numerical Methods in Biomedical Engineering 34, 1–14.

Garot, P., Iriart, X., Aminian, A., Kefer, J., Freixa, X., Cruz-Gonzalez, I., Berti, S., Rosseel, L., Ibrahim, R., Korsholm, K., Odenstedt, J., Nielsen-Kudsk, J.E., Saw, J., Sondergaard, L., De Backer, O., 2020. Value of FEops HEARTguide patient-specific computational simulations in the planning of left atrial appendage closure with the Amplatzer Amulet closure device: Rationale and design of the PREDICT-LAA study. Open Heart 7, 1–5.

Goo, H.W., Park, S.J., Yoo, S.J., 2020. Advanced medical use of three-dimensional imaging in congenital heart disease: Augmented reality, mixed reality, virtual reality, and three-dimensional printing. Korean journal of radiology 21, 133–145.

Hascoet, S., Smolka, G., Bagate, F., Guihaire, J., Potier, A., Hadeed, K., Lavie-Badie, Y., Bouvaist, H., Dauphin, C., Bauer, F., Nejjari, M., Pillière, R., Brochet, E., Mangin, L., Bonnet, G., Ciobotaru, V., Leurent, G., Hammoudi, N., Aminian, A., Karsenty, C., Spaulding, C., Armero, S., Collet, F., Champagnac, D., Ternacle, J., Kloeckner, M., Gerardin, B., Isorni, M.A., 2018. Multimodality imaging guidance for percutaneous paravalvular leak closure: Insights from the multi-centre ffpp register. Archives of Cardiovascular Diseases 111.

Karagodin, I., Addetia, K., Singh, A., Dow, A., Rivera, L., DeCara, J.M., Soulat-Dufour, L., Yamat, M., Kruse, E., Shah, A.P., Mor-Avi, V., Lang, R.M., 2020. Improved Delineation of Cardiac Pathology Using a Novel Three-Dimensional Echocardiographic Tissue Transparency Tool. Journal of the American Society of Echocardiography : official publication of the American Society of Echocardiography 33, 1316–1323.

Kikinis, R., Pieper, S.D., Vosburgh, K.G., 2014. 3d slicer: A platform for subject-specific image analysis, visualization, and clinical support. Intraoperative Imaging and Image-Guided Therapy.

Kohli, K., Wei, Z.A., Sadri, V., Easley, T.F., Pierce, E.L., Zhang, Y.N., Wang, D.D., Greenbaum, A.B., Lisko, J.C., Khan, J.M., Lederman, R.J., Blanke, P., Oshinski, J.N., Babaliaros, V., Yoganathan, A.P., 2020. Framework for Planning TMVR using 3-D Imaging, In Silico Modeling, and Virtual Reality. Structural Heart 0, 1–6.

Masci, A., Alessandrini, M., Forti, D., Menghini, F., Dedé, L., Tomasi, C., Quarteroni, A., Corsi, C., 2020. A Proof of concept for computational fluid dynamic analysis of the left atrium in atrial fibrillation on a patient-specific basis. Journal of Biomechanical Engineering 142, 1–11.

Medina, E., Aguado, A.M., Mill, J., Freixa, X., Arzamendi, D., Yagüe, C., Camara, O., 2020. VRIDAA: Virtual Reality Platform for Training and Planning Implantations of Occluder Devices in Left Atrial Appendages, in: Kozlíková, B., Krone, M., Smit, N., Nieselt, K., Raidou, R.G. (Eds.), Eurographics Workshop on Visual Computing for Biology and Medicine, The Eurographics Association.

Mendez, A., Hussain, T., Hosseinpour, A.R., Valverde, I., 2018. Virtual reality for preoperative planning in large ventricular septal defects. European Heart Journal 40, 1092–1092.

Mill, J., Olivares, A.L., Arzamendi, D., Agudelo, V., Regueiro, A., Camara, O., Freixa, X., 2020. Impact of flow dynamics on device-related thrombosis after left atrial appendage occlusion. Canadian Journal of Cardiology 36.

Mill, J., Olivares, A.L., Noailly, J., Freixa, X., Camara, O., 2019. Optimal Boundary Conditions in Fluid Simulations for Predicting Occluder-Related Thrombus Formation in the Left Atria, in: 6th International Conference on Computational and Mathematical Biomedical Engineering – CMBE2019, Sendai, Japan. pp. 256–259.

Naci, H., Salcher-Konrad, M., Mcguire, A., Berger, F., Kuehne, T., Goubergrits, L., Muthurangu, V., Wilson, B., Kelm, M., 2019. Impact of predictive medicine on therapeutic decision making: a randomized controlled trial in congenital heart disease. npj Digital Medicine 2, 1–9.

Nam, H., Herz, C., Lasso, A., Drouin, S., Posada, A., Morray, B., O’Byrne, M., Paniagua, B., Joffe, D., Mackensen, B., Rogers, L., Fichtinger, G., Jolley, M., 2020. Simulation of transcatheter atrial and ventricular septal defect device closure within three-dimensional echocardiography–derived heart models on screen and in virtual reality. Journal of the American Society of Echocardiography 33.

Narang, A., Hitschrich, N., Mor-Avi, V., Schreckenberg, M., Schummers, G., Tiemann, K., Hitschrich, D., Sodian, R., Addetia, K., Lang, R.M., Mumm, B., 2020. Virtual Reality Analysis of Three-Dimensional Echocardiographic and Cardiac Computed Tomographic Data Sets. Journal of the American Society of Echocardiography 33, 1306–1315.

Otani, T., Al-Issa, A., Pourmorteza, A., McEigh, E., Wada, S., Ashikaga, H., 2016. A Computational Framework for Personalized Blood Flow Analysis in the Human Left Atrium. Annals of biomedical engineering 44, 3284–3294.

Ribeiro, J.M., Hokken, T., Astudillo, P., Jan Nuis, R., de Backer, O., Rocatello, G., Daemen, J., M Van Mieghem, N., Cummins, P., de Beule, M., Lumens, J., Bruining, N., PT de Jaegere, P., 2020. Artificial intelligence and advanced computer modelling in transcatheter interventions for structural heart disease - implications for clinical practice. Digital Health Virtual Journal, 1–12.

Salavitabar, A., Figueroa, C.A., Lu, J.C., Owens, S.T., Axelrod, D.M., Zampi, J.D., 2020. Emerging 3D technologies and applications within congenital heart disease: teach, predict, plan and guide. Future Cardiology 0.

Sanon, S., Lim, D.S., 2019. Update on left atrial appendage occlusion. Cardiac interventions today 13.

Saw, J., Fahmy, P., Spencer, R., Prakash, R., McLaughlin, P., Nicolaou, S., Tsang, M., 2016. Comparing measurements of ct Angiography, TEE, and fluoroscopy of the left atrial appendage for percutaneous closure. Journal of Cardiovascular Electrophysiology 27. 1011.1669v3.

Southworth, M.K., Silva, J.R., Silva, J.N., 2020. Use of extended realities in cardiology. Trends in Cardiovascular Medicine 30, 143–148.

Tandon, A., Burkhardt, B.E., Batsis, M., Zellers, T.M., Velasco Forte, M.N., Valverde, I., McMahan, R.P., Guleserian, K.J., Greil, G.F., Hussain, T., 2019. Sinus Venosus Defects: Anatomic Variants and Transcatheter Closure Feasibility Using Virtual Reality Planning. JACC: Cardiovascular Imaging 12, 921–924.

Vukicevic, M., Mosadegh, B., Min, J.K., Little, S.H., 2017. Cardiac 3D Printing and its Future Directions. JACC: Cardiovascular Imaging 10, 171–184.

Wang, D.D., Geske, J., Choi, A.D., Khalique, O., Lee, J., Atianzar, K., Wu, I., Blanke, P., Gafoor, S., Cavalcante, J.L., 2018a. Navigating a Career in Structural Heart Disease Interventional Imaging. JACC: Cardiovascular Imaging 11, 1928–1930.

Wang, D.D., Gheewala, N., Shah, R., Levin, D., Myers, E., Rollet, M., O’Neill, W.W., 2018b. Three-Dimensional Printing for Planning of Structural Heart Interventions. Interventional Cardiology Clinics 7, 415–423.

Wang, D.D., Qian, Z., Vukicevic, M., Engelhardt, S., Kheradvar, A., Zhang, C., Little, S.H., Verjans, J., Comaniciu, D., O’Neill, W.W., Vannan, M.A., 2021. 3D Printing, Computational Modeling, and Artificial Intelligence for Structural Heart Disease. JACC. Cardiovascular imaging 14, 41–60.

Zbroński, K., Rymuza, B., Scisło, P., Kochman, J., Huczek, Z., 2018. Augmented reality in left atrial appendage occlusion. Kardiologia polska 76, 212.

