## Supplemental SUS questionnaire for "Domain expert evaluation of advanced visual computing solutions for the planning of left atrial appendage occluder interventions"

### SUPPLEMENTARY MATERIAL

#### Appendix A: SUS questionnaire

1. I think that I would like to use this system frequently

| Strongly agree | Agree | More or less agree | Undecided | More or less disagree | Disagree | Strongly disagree |
|---|---|---|---|---|---|---|

2. I found the system unnecessarily complex

| Strongly agree | Agree | More or less agree | Undecided | More or less disagree | Disagree | Strongly disagree |
|---|---|---|---|---|---|---|

3. I thought the system was easy to use

| Strongly agree | Agree | More or less agree | Undecided | More or less disagree | Disagree | Strongly disagree |
|---|---|---|---|---|---|---|

4. I think that I would need the support to use this system

| Strongly agree | Agree | More or less agree | Undecided | More or less disagree | Disagree | Strongly disagree |
|---|---|---|---|---|---|---|

5. I found the various functions in this system were well integrated

| Strongly agree | Agree | More or less agree | Undecided | More or less disagree | Disagree | Strongly disagree |
|---|---|---|---|---|---|---|

6. I thought there was too much inconsistency in this system

| Strongly agree | Agree | More or less agree | Undecided | More or less disagree | Disagree | Strongly disagree |
|---|---|---|---|---|---|---|

7. I would imagine that most people would learn to use this system very quickly

| Strongly agree | Agree | More or less agree | Undecided | More or less disagree | Disagree | Strongly disagree |
|---|---|---|---|---|---|---|

8. I found the system very cumbersome to use

| Strongly agree | Agree | More or less agree | Undecided | More or less disagree | Disagree | Strongly disagree |
|---|---|---|---|---|---|---|

9. I felt very confident using the system

| Strongly agree | Agree | More or less agree | Undecided | More or less disagree | Disagree | Strongly disagree |
|---|---|---|---|---|---|---|

10. I needed to learn a lot of things before I could get going with this system

| Strongly agree | Agree | More or less agree | Undecided | More or less disagree | Disagree | Strongly disagree |
|---|---|---|---|---|---|---|

### Appendix B: SUS questionnaire results

| | VIDAA | VRIDAA | 3D Printing | Simulations |
|---|---|---|---|---|
| Q1 | 5.0/6.0 | 3.2/6.0 | 4.5/6.0 | 4.5/6.0 |
| Q2 | **5.3/6.0** | 4.7/6.0 | 4.8/6.0 | 3.0/6.0 |
| Q3 | **5.3/6.0** | **5.1/6.0** | **5.5/6.0** | **2.8/6.0** |
| Q4 | 4.0/6.0 | 3.1/6.0 | **5.2/6.0** | **2.5/6.0** |
| Q5 | 4.3/6.0 | **2.8/6.0** | **2.8/6.0** | 4.6/6.0 |
| Q6 | 4.6/6.0 | 4.0/6.0 | 3.0/6.0 | 4.5/6.0 |
| Q7 | **5.5/6.0** | 4.7/6.0 | **5.6/6.0** | 3.0/6.0 |
| Q8 | 5.0/6.0 | 4.7/6.0 | **5.3/6.0** | 3.2/6.0 |
| Q9 | 5.0/6.0 | 4.7/6.0 | 4.0/6.0 | 4.0/6.0 |
| Q10 | 4.0/6.0 | 4.3/6.0 | 5.0/6.0 | 4.2/6.0 |

***Table 1:*** *The SUS average results obtained in each the question from the 6 participants. Maximum value obtained per question was 6.*
